# Increased CRTH2 Expression in Postural Orthostatic Tachycardia Syndrome

**DOI:** 10.1101/2024.11.18.24317517

**Authors:** Julian Achemdey, Jose Manuel Martin, Hala Abdallah, Mian Li, Victor E. Nava, Hasan Abdallah

## Abstract

**Purpose:** The exact etiology of Postural Orthostatic Tachycardia Syndrome (POTS) is still unknown but is likely heterogeneous and multifaceted, involving autoimmune, neuropathic, hypovolemic, and hyperadrenergic pathways. Recently, more research has been devoted to understanding POTS’s underlying immune etiology. We investigated the potential association between elevated levels of prostaglandin chemoattractant receptors (CRTH2) expressed on T-cells and the clinical manifestations observed in patients with POTS.

**Methods:** We performed immunophenotyping using flow cytometry on 20 consecutive patients with POTS confirmed by tilt table testing and compared our findings with healthy controls.

**Results:** POTS patients demonstrated an increase in CRTH2 levels on (CD8+) T cells and CD8+/CD45RO+ T cells compared to controls.

**Conclusions:** CRTH2 is expressed significantly on TH2 and CD8+ cells in POTS patients compared with controls. Since CRTH2 mediates the action of mast cell degranulation mediators such as PGD2 on TH2 cells it results in the release of inflammatory cytokines and interleukins that cause vasodilation and cellular inflammation with capillary leak, leading to reduced effective intravascular volume and reduced venous return, which are known as potential mechanisms in the pathophysiology of POTS, we conclude that CRTH2 plays an important role in the pathophysiology of POTS.

## INTRODUCTION

Postural orthostatic tachycardia syndrome (POTS) is a heterogeneous clinical condition characterized by sustained and excessive sinus tachycardia upon standing, without orthostatic hypotension, and with chronic symptoms of orthostatic intolerance [1] POTS primarily affects young females and is estimated to impact approximately 0.2% to 1% of the general population in the United States, equivalent to about 1-3 million individuals [2] Diagnostic criteria include a sustained increase in heart rate of more than 30 beats per minute (BPM) in adults and more than 40 BPM in younger individuals within 10 minutes of assuming an upright position, along with symptoms such as lightheadedness, palpitations, and tremulousness [2]. Additional manifestations such as headaches, chronic fatigue, and exercise intolerance may also be present. There are also patients who have additional symptoms that are not typical of POTS such as abdominal bloating, diarrhea, skin itching, and bronchospasm, which are thought to be due to mast cell degranulation [3]. Mast cell degranulation releases various mediators, including prostaglandin D2 (PGD2) that binds its cognate receptor CRTH2 (chemoattractant receptor-homologous molecule expressed on T helper type 2 cells, also known as PGD2 receptor or CD294) expressed mainly on Type 2 CD4+ helper T cells and a subset of basophils, eosinophils and CD8+ cytotoxic T cells (Type 2 cytotoxic T cells) [4]. Activation of CRTH2 upon binding of ligands is essential in allergy [5]. However, its role in POTS has not been documented in PubMed. Herein we report elevated levels of CRTH2 expression on cytotoxic lymphocyte subsets from POTS patients using flow cytometry and discuss potential clinical ramifications of the finding.

## METHODS

The patient population consisted of patients referred to the autonomic clinic at the Children’s Heart Institute - Johns Hopkins Medicine, in Herndon, Virginia for evaluation of POTS and to rule out any other underlying or associated condition. Institutional Review Board approvals from the Johns Hopkins Review Board and consent from all patients for the use of deidentified data were obtained. Researchers in the field will have access to tabular versions of the deidentified data used for this study upon request. All patients were evaluated by the principal investigator (HIA). Data collection for this study was conducted prior to the onset of the COVID-19 pandemic. Patients were included in the study if they did not have any other diagnoses that could present as POTS, such as cardiomyopathy, significant intracardiac shunting, systemic inflammatory diseases, adrenal failure, hyperthyroidism, concurrent viral illness, or orthostatic symptoms of less than 6 months duration. A detailed medical history with comprehensive symptom assessment, physical examination, a 12-lead ECG, Echocardiogram, 24-Hour Holter, and laboratory testing to exclude anemia, acute inflammatory, autoimmune diseases, and infectious illness, were performed. Complete blood cell count with white blood cell differential, erythrocyte sedimentation rate, C-reactive protein, comprehensive chemistry panel, thyroid panel, cortisol AM and PM levels, aldosterone level, and urine analysis were also conducted. In addition, 10-color flow cytometry (BD FACS Lyric), was performed on peripheral blood with the following panel. The diagnosis of POTS was based on positive tilt table testing that confirmed a sustained increase in heart rate (HR) ≥ 30 beats per minute (BPM) in patients older than 18 years, or HR ≥ 40 BPM in patients younger than 18 years, in the absence of sustained orthostatic hypotension, i.e. a 20 mmHg drop of systolic blood pressure (BP) or 10 mmHg drop in diastolic BP within 10 minutes of passive tilt from the supine to the upright position based on previously published guidelines [3]. This study employed a retrospective design to review flow cytometry results and tilt table test results of a cohort of 20 patients confirmed as having POTS based on the above-established criteria. These results were compared with data obtained from 20 control subjects without symptoms of POTS and analyzed by Fisher’s t-test. Patients who, apart from experiencing typical POTS symptoms, also reported symptoms less often associated with typical POTS (e.g., Gastrointestinal symptoms, “allergy,” skin flushing, rash/urticaria), underwent laboratory measurements of common mast cell mediators and metabolites, including the following: plasma tryptase, either plasma prostaglandin D2 or 24-hour urine prostaglandin 11-β-PGF 2alpha or both, plasma histamine, and urine methylhistamine. Timing of laboratory measurements was based on patient convenience and was not specifically associated with symptom outbreak. In some cases, complete laboratory studies could not be obtained due to insurance coverage constraints. Descriptive statistics were utilized for the statistical analysis, including measures such as mean, standard deviation, minimum, maximum, and percentiles. P-values were calculated to assess the significance of intergroup differences, using also Joint Analysis method. The investigation also evaluated the frequency and prevalence of reported symptoms in POTS patients to provide insights into symptom distribution in relation to biomarker values.

## RESULTS

The study population comprised 20 patients with POTS, aged 13–52 years (15 females [75%] and 5 males [25%]), matched with 20 healthy controls, aged 12–49 years (15 females [75%] and 5 males [25%]), who did not have any orthostatic symptoms. Patients with POTS reported a markedly higher prevalence of symptoms across various domains compared to controls. Specifically, POTS patients frequently experienced brain fog (95% vs. 0%), dizziness (75% vs. 0%), palpitations (70% vs. 0%), fatigue (80% vs. 0%), throbbing headaches (45% vs. 0%), and depression/anxiety (40% vs. 0%). Additionally, presyncope (35% vs. 0%), syncope (55% vs. 0%), blood pooling in extremities (50% vs. 0%), allergies (15% vs. 0%), nasal congestion (20% vs. 0%), pruritic skin rash (20% vs. 0%), abdominal bloating (30% vs. 0%), diarrhea (15% vs. 0%), nausea (35% vs. 0%), and abdominal pain (30% vs. 0%) were significantly more prevalent among POTS patients compared to controls (Table 1).

**Table 1:**
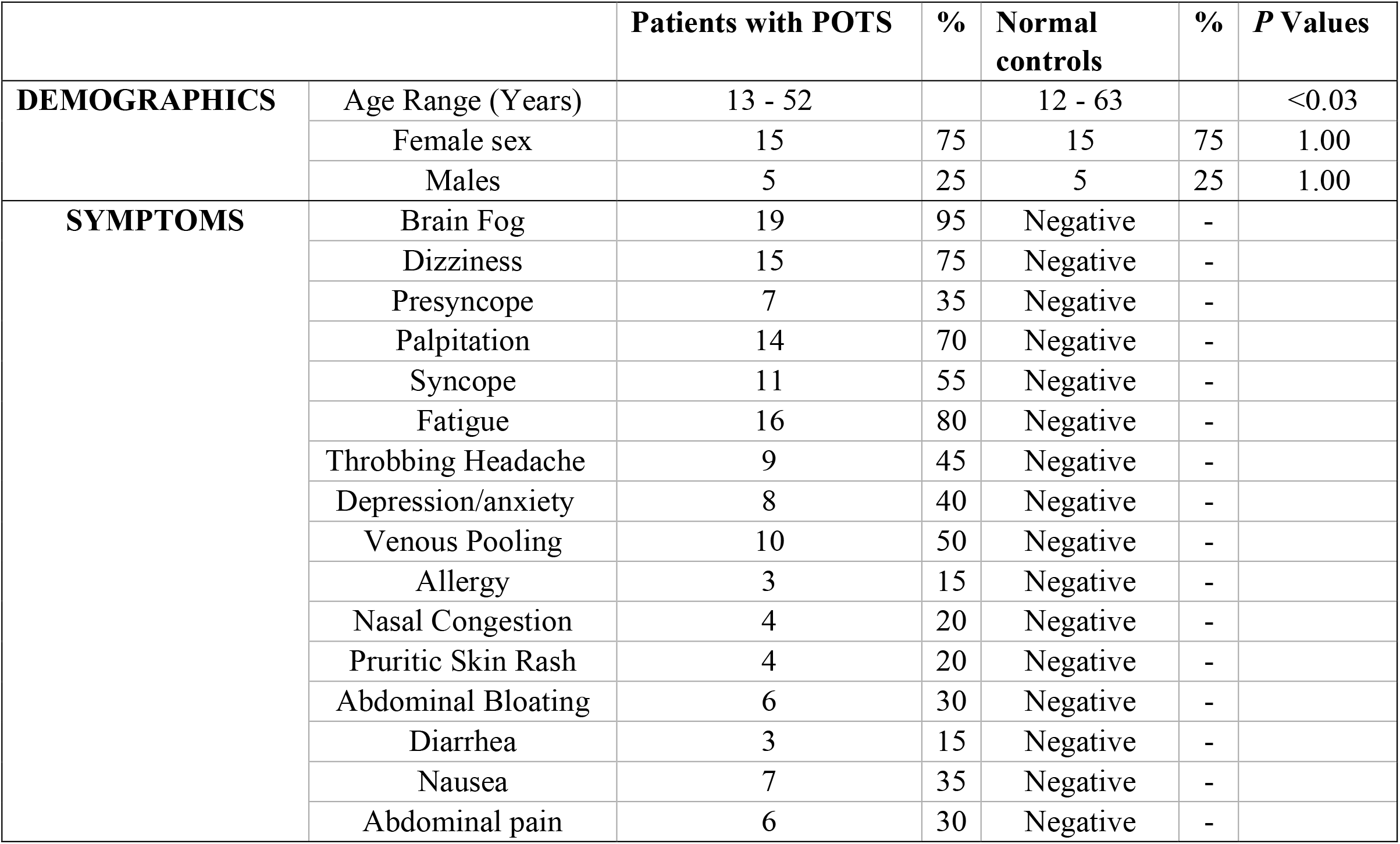
Clinical characteristics of subject.

In agreement with Gunning et al [6] suggesting an autoinflammatory etiology for POTS, we detected alteration in the lymphocyte’s subsets in comparison with controls. Notably, POTS patients showed an increased percentage of CD3-positive T-lymphocytes co-expressing CRTH2 and CD4 (helper T cells) and CD8 (cytotoxic T cells) among other markers (Table 2 and data not shown).

**Table 2:**
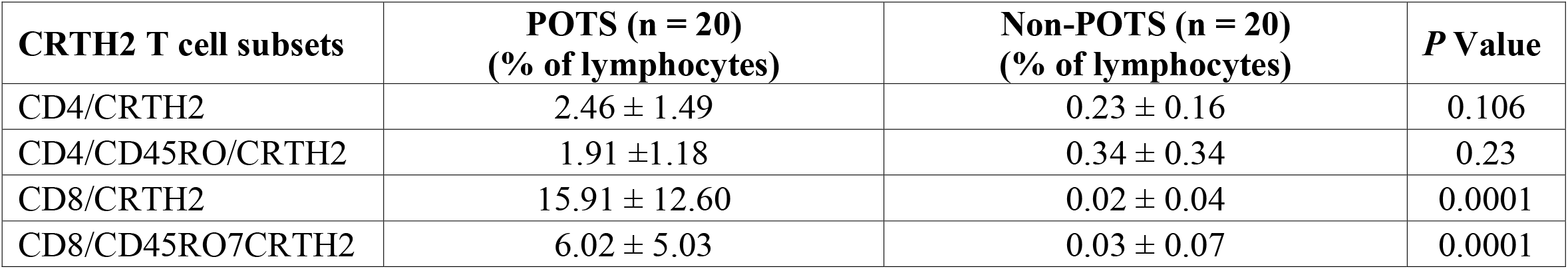
Flow cytometry quantification of T cell subsets expressing CRTH2.

Specifically, marked differences in T lymphocytes co-expressing CD4/CRTH2, CD4/CD45RO/CRTH2, CD8/CRTH2, and/or CD8/CD45RO/CRTH2 were observed in POTS patients relative to non-POTS controls. Interestingly, a statistically significantly higher percentage of T cytotoxic lymphocytes was detected in POTS. The differences between POTS and control subjects in CD8/CRTH2 (15.91 ± 12.60 vs. 0.02 ± 0.04) and CD8/CD45RO/CRTH2 (6.02 ± 5.03 vs. 0.03 ± 0.07) showed a P = 0.0001 for both sets. However, the differences in CD4/CRTH2 (2.46 ± 1.49 vs. 0.23 ± 0.16) and CD4/CD45RO/CRTH2 (1.91 ± 1.18 vs. 0.34 ± 0.34) were not statistically significant (P = 0.106 and 0.23 respectively) from controls, which was also the case for all other markers in the panel (data not shown).

Besides flow cytometry, which was performed with the same panel in all subjects, other laboratory tests performed were based on the provider’s discretion, individual symptoms, and insurance coverage. None of the patients had abnormal thyroid function (data not shown). Although some differences were seen in absolute eosinophil counts, serum PGD2, tryptase and serum N-Methylhistamine, none of these differences were statistically significant (Table 3).

**Table 3:**
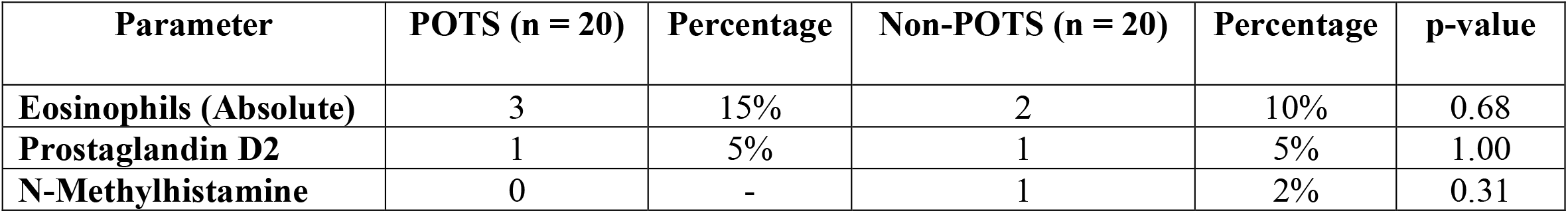
Elevated values of Eosinophils, Prostaglandin D2, and N-Methylhistamine in selected patients.

In addition, antinuclear antibodies were mildly elevated (41 IU/mL) in 1/20 POTS patients non-specifically.

## DISCUSSION

Even though a unified physiopathological mechanism for POTS is not established, various coexisting processes have been identified in primary cases, including autonomic nervous system dysregulation, inappropriate peripheral blood flow control and autoimmune disorders [7]. Furthermore, the well-known overlap between immunologic conditions such as MACS and POTS (seen in up to 30% of patients) highlights the possibility of immune dysregulation as a mediator or amplifier of orthostatic hypotension [8].

Our study reveals a statistically significant increase in CRTH2 levels on a subset of cytotoxic lymphocytes from POTS patients. Specifically, CRTH2 was elevated in CD8+ and CD8+/CD45RO+ memory T cells compared to age- and sex-matched controls. In contrast, POTS patients did not show a significant CRTH2 elevation on helper CD4+ T cells, which are typically linked to type 2 immune responses. CRTH2 has been identified as a crucial factor in MCAS, though its role in POTS remains to be established.

Prostaglandin D2 (PGD2), released by mast cells during allergen exposure, is crucial in inflammation related to allergies and asthma. Its effects are mediated through several receptors, including activation of CRTH2, a G-protein-coupled receptor found on immune cells like lymphocytes, eosinophils, basophils, monocytes, and mast cells [9]. Upon binding PGD2 released by mast cells induces a Th2-like cytokine response characterized by IL-4 and IL-13 release [10], capable of inducing inflammatory-associated symptoms, such as brain fog, which was reported in 95% of POTS patients. Interestingly, we failed to detect differences in PGD2 levels between POTS and control patients. These results suggest the possibility of a CRTH2-mediated response even with ordinary PGD2 levels in POTS. Refer to Figure 1.

**Figure 1:**
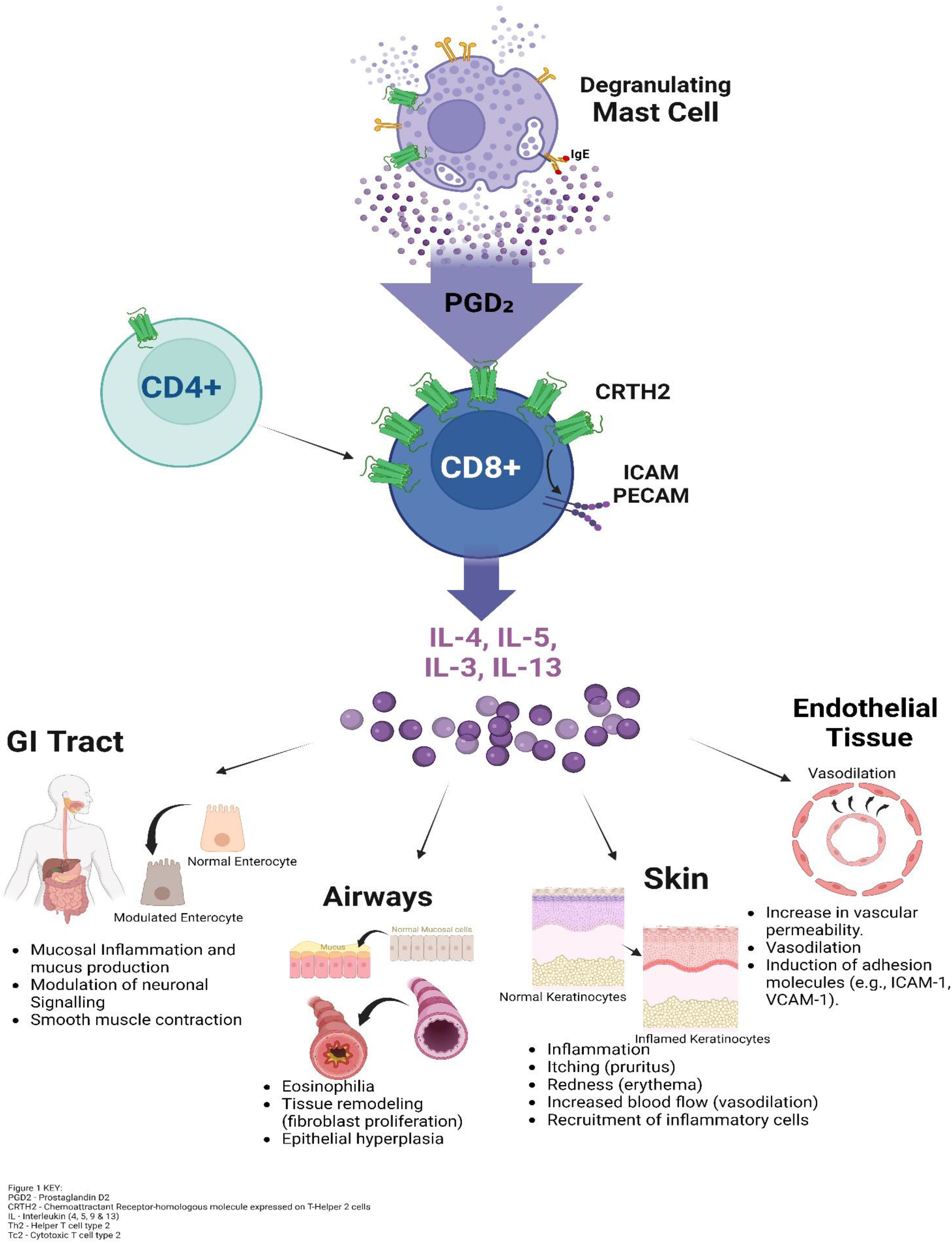
Activation of T cells by mast cells and prostaglandin D2 (PGD2), leading to cytokine release (IL-4, IL-5, IL-3, IL-13) that contributes to inflammation in the gastrointestinal tract, airways, skin, and endothelial tissues.

As in asthma, Type 2 cytotoxic CD8+ T cells (Tc2 cells), distinct from Tc1 cells, might play a significant role in POTS [11]. While Tc1 cells produce IFN-γ for direct pathogen elimination, Tc2 cells release cytokines (such as IL-4, IL-5 and IL-13) associated with type 2 immune responses, including allergies and parasite-induced immune response [12]. Although no evidence of CD4+ T cells direct involvement in POTS was found, an indirect effect amplifying a Tc2 response is possible. Further research is needed to confirm this hypothesis and to elucidate the interactions between Tc2 cells and mast cells in POTS.

Furthermore, it is important to consider the association of POTS with MCAS, as it significantly impacts therapeutic approaches. Patients with elevated CRTH2 in our study presented a clinical picture distinct from typical POTS, characterized by symptoms such as palpitations, dizziness, flushing, and brain fog. These symptoms were often accompanied by manifestations of mast cell activation, such as nausea, abdominal bloating, flushing, and allergic symptoms, in agreement with previous reports [3]. While there were variations in absolute eosinophil counts, serum PGD2, tryptase, and serum N-Methylhistamine levels between POTS and controls, none of these differences reached statistical significance. Further studies with larger sample size and more frequent analyte monitoring may provide additional insights into the biochemical profiles associated with POTS.

In summary, our findings suggest that CRTH2 may serve as both a valuable biomarker and a promising therapeutic target for future clinical management strategies in POTS. This is particularly relevant for POTS patients presenting with symptoms of MCAS. Targeting the PGD2/CRTH2 signaling pathway, potentially through agents like fevipiprant and setipiprant, [13] [14] could offer new therapeutic avenues yielding symptomatic relief for this challenging syndrome.

## Data Availability

Data will be made available upon request.

## Acknowledgements

We gratefully acknowledge the participants who generously contributed to this study. We also acknowledge the valuable insights and feedback from colleagues and reviewers that have contributed to the refinement of this manuscript.

## Authorship contribution

Dr. Hasan Abdallah, the principal investigator, in conjunction with Dr. Hala Abdallah, contributed to patient care and manuscript review. Julian Achemdey managed data collection, analysis, coordination of author contributions, and prepared figures and references. Dr. Jose Manuel Martin analyzed data and conducted the literature search. Dr. Mian Li analyzed data and edited the manuscript. Dr. Victor E. Nava designed and analyzed flow cytometry data and edited the manuscript.

